# Maternal and perinatal characteristics and outcomes of pregnancies complicated with COVID-19 in Kuwait

**DOI:** 10.1101/2020.07.10.20150623

**Authors:** Amal Ayed, Alia Embaireeg, Asma Benawath, Wadha Al-Fouzan, Majdeda Hammoud, Monif Al-Hathal, Abeer Alzaidai, Mariam Ayed

**Affiliations:** Obstetrics and Gynecology Department, Farwaniya Hospital, Kuwait; Paediatric Department, Farwaniya Hospital, Kuwait; Obstetrics and Gynecology Department, Maternity Hospital, Kuwait; Department of Microbiology, Faculty of Medicine, Kuwait University; Paediatrics Department, Faculty of Medicine, Kuwait University; Neonatal Department, Maternity Hospital, Kuwait; Obstetrics and Gynaecology Department, Al-Adan Hospital, Kuwait; Neonatal Department, Farwaniya Hospital, Kuwait

**Keywords:** SARS-CoV-2, COVID-19, Pregnant women, Neonates, Clinical features

## Abstract

**Background:** In late December of 2019, a novel coronavirus (SARS-CoV-2) was identified in the Chinese city Wuhan among a cluster of pneumonia patients. While it is known that pregnant women have reduced immunity and they are at risk for COVID-19 infection during the current pandemic, it is not clear if the disease manifestation would be different in pregnant women from non-pregnant women.

**Objectives:** To describe the maternal and neonatal clinical features as well as outcome of pregnancies complicated with SARS-CoV-2 infection.

**Methods:** In this retrospective national-based study, we analyzed the medical records of all SARS-CoV-2 positive pregnant patients and their neonates who were admitted to New-Jahra Hospital, Kuwait, between March 15^th^ 2020 and May 31^st^ 2020. The outcomes of pregnancies were assessed until the end date of follow-up (June 15^th^ 2020).

**Results:** A total of 185 pregnant women were enrolled with a median age of 31 years (interquartile range, IQR: 27.5-34), and median gestational age at diagnosis was 29 weeks (IQR: 18-34). The majority (88%) of the patients had mild symptoms, with fever (58%) being the most common presenting symptom followed by cough (50.6%). During the study period, 141 (76.2%) patients continued their pregnancy, 3 (1.6%) had a miscarriage, 1 (0.5%) had intrauterine fetal death and only 2 (1.1%) patients developed severe pneumonia and required intensive care. Most of the neonates were asymptomatic, and only 2 (5%) of them tested positive on day 5 by nasopharyngeal swab testing.

**Conclusion:** Pregnant women do not appear to be at higher risk to the COVID-19 than the general population. The clinical features of pregnant women with SARS-CoV-2 infection were similar to those of the general population having SARS-CoV-2 infection. Favorable maternal and neonatal outcomes reinforce the existing evidence and may guide healthcare professionals in the management of pregnancies complicated with SARS-CoV-2 infection.

## Introduction

In late December of 2019, a novel coronavirus (SARS-CoV-2) that emerged in the Chinese city Wuhan was identified among a cluster of pneumonia cases [1]. The outbreak of coronavirus disease (COVID-19) rapidly spread to become a global pandemic as declared by the World Health Organization in March 2020 [1-3]. This advent and remarkable widespread SARS-CoV-2 reflects a new public health crisis [4].

SARS-CoV-2 is a member of the Coronavirus family, and other pathogens from this family have inflicted a range of viral infections, including Middle East Respiratory Syndrome (MERS) and Severe Acute Respiratory Syndrome (SARS) [5]. The main route of transmission is through respiratory droplets and direct contact. While the spectrum of the disease severity ranges from mild to critical, most are mild. Most of the fatal cases occurred in patients with advanced age or underlying comorbidities [6]. However, to this date, the SARS-CoV-2 mortality rate is greater than MERS and SARS combined and have a global Case Fatality Rate of about 6.4% [1].

Although it is known that pregnant women have a reduced immunity [5], and they are at risk for COVID-19 infection during the current pandemic, it is not clear how the disease manifestation would be different in pregnant women from non-pregnant women. It is known that the body’s immune system and response to viral infections might be changed as a result of pregnancy, and this explains the cause of more severe symptoms [7]. However, there is currently no evidence that pregnant women are more susceptible to SARS-CoV-2 than non-pregnant women [8 – 11]]. They may present with severe diseases that include hypoxia, hypotension, electrolyte disturbances, placental hypo perfusion, which can provoke fetal distress, preterm labor, miscarriage, or fetal death [12 – 18].

Preliminary evidence from the diseases caused by H1N1 virus (H1N1 influenza) and other coronaviruses (SARS and MERS) have suggested that pregnant women are more likely to have severe manifestations of the disease, morbidity, and mortality as compared to non-pregnant women [19 – 22]. Moreover, some studies have shown an association between the risk of these critical illnesses and later stages of pregnancy [23 – 25].

To date, information about the different routes (transplacental, perinatal, intrapartum and postnatal) and risks of transmission of COVID-19 to newborn infants is based on case series [13]. In consideration of the fact that SARS-CoV-2 is a respiratory virus, the risk of vertical transmission is supposed to be very low or absent. This supposition is supported by the evidence that there was no vertical transmission detected in other members of the coronavirus family: SARS and MERS [19].

The risk of perinatal transmission of SARS-CoV-2 is unknown. Additionally, since the risk of postnatal transmission remains to be elucidated, it is debatable whether the newborn should be immediately separated from the COVID-19 positive mother [8]. A study comprised of outcomes of 7 newborns necessitates the separation of newborns to avoid potential threats of SARS-CoV-2 infection [26]. In another study, contrary to the previous one, it is concluded that separation may not prevent infection, and early separation might increase neonatal risk of pneumonia [27]. However, the severity of postnatally acquired disease in the newborn is unknown. A case series of 10 COVID-19 negative neonates born to COVID-19 positive mothers reported fetal distress, premature labor, respiratory distress, thrombocytopenia accompanied by abnormal liver function, and even death among neonates [12]. This may indicate a possible association, but not necessarily a causal effect.

We believe that the information on outcomes of such pregnancies could continuously update guidance and management procedures for COVID-19 positive pregnant mothers and their neonates. Therefore, the study aimed at describing the maternal as well as neonatal clinical features and outcome of pregnancies complicated with SARS-CoV-2 infection.

## Methods

### Study Design

We performed a retrospective national-based analysis of medical records from all SARS-CoV-19 positive pregnant patients and their neonates who were admitted to New-Jahra Hospital, Kuwait between March 15^th^, 2020 and May 31^st^, 2020, and the outcomes were assessed till the end date of follow-up (June 15^th^, 2020). New Jahra hospital was designated as a national hospital for the management of COVID-19 infected pregnant patients and neonates born to infected mothers in Kuwait. As part of Kuwait’s national COVID-19 policy, all pregnant women were tested if they had SARS-CoV-2 symptoms or had been in contact with infected patients. Moreover, all confirmed COVID-19 positive pregnant women were admitted to the maternity department for at least 14 days or till resolution of symptoms. Patients management was based on local and international guideline protocols (<u>Appendix 1</u>).

The study was approved by the Ethics Committee of the Ministry of Health of Kuwait (2020/1420)

### Patient selection criteria

We enrolled all pregnant women admitted to the maternity department due to SARS-CoV-2 infection, as confirmed by real-time reverse transcriptase-polymerase chain reaction (RT-PCR) assay of nasopharyngeal swab specimens (Cobas 6800 Systems, Roche, Switzerland) / (Taq Path, Thermo-Fisher Scientific, USA). Patients who had equivocal or negative testing results were excluded from the study.

### Assessment of neonates

The neonatal team assessed neonates after delivery. They were isolated immediately and admitted to a designated neonatal intensive care unit. All neonates born to SARS-CoV-2 positive mothers were admitted to a negative pressure isolated single room, cared by health care providers wearing full personal protective equipment, and were fed artificial formula. Moreover, nasopharyngeal swab testing was performed for all neonates on day 5 and then on day 14.

### Data collection

Maternal and neonatal data were extracted from the electronic medical records or medical charts using a standardized data collection sheet. For maternal data, we recorded the information on gestational age at diagnosis of SARS-CoV-2, parity (nulliparous or multiparous), pre-existing medical condition (i.e., including hypertension, diabetes, bronchial asthma, hypothyroidism, and autoimmune diseases) and presenting symptoms prior to admissions, such as fever, sore throat/ nasal or sinus congestion, cough/chest pain, vomiting/diarrhea, and fatigue/myalgia).

Information on current pregnancy was also collected, including gestational diabetes, pregnancy-induced hypertension, pre-eclampsia, and multiple pregnancies. We collected the laboratory and chest x-ray results upon admission to the hospital. Information on patient-specific therapies, such as antenatal steroids for lung maturation, administration of antibiotics, antivirals, and thromboprophylaxis (i.e. low molecular weight heparin was also collected. For patients admitted to intensive care unit, we obtained data on maximum respiratory support (i.e. intubation and mechanical ventilation), acute respiratory distress syndrome (ARDS) as defined according to the Berlin criteria and duration of ventilation [28]

For neonatal data, we collected gestational age at delivery, mode of delivery, birth weight, 1 and 5 minutes Apgar score, and the results of nasopharngeal swabs. Data entry and quality were checked and reviewed independently by two investigators (AE and AB).

We defined gestational age as the number of weeks calculated according to the last normal menstrual period or expected date of delivery according to an early ultrasound scan. Gestational diabetes was defined as high blood sugar (glucose) using the Oral Glucose Tolerance Test as a screening method (fasting blood sugar ≥ 5.1 mmol/L or 1 hour postprandial ≥10 mmol/L or 2 hours postprandial ≥8.5 mmol/L) that developed during pregnancy and usually disappeared after giving birth. Preeclampsia was defined as new onset of hypertension (over 140 mmHg systolic or over 90 mmHg diastolic) after 20 weeks of pregnancy and significant proteinuria. Pregnancy-induced hypertension (PIH) was defined as a new hypertension presenting after 20 weeks of pregnancy without significant proteinuria.

### Statistical analysis

We reported continuous variables as mean ± standard deviation (SD) or median (interquartile range [IQR]) and categorical variables as number and percentage. Statistical analysis was performed using STATA/IC 14 software (STATA Corp, College Station, Texas).

## Results

During the study period, 185 pregnant women were identified with confirmed SARS-CoV-2 infection with a median age of patients was 31 years (IQR: 27.5-34), and 19.5% were nulliparous. The majority of patients were healthy, 19 (10.3%) patients had gestational diabetes, and 4 (2.3%) had PIH. The median gestational age at diagnosis of SARS-CoV-2 was 29 weeks (IQR: 18 to 34), and half of the women were in their 3^rd^ trimester. Additionally, the majority of the patients (88.6%) were symptomatic. The most common symptom was fever in 105 (58%) patients followed by cough in 90 (50.6%) patients, and the median duration of symptoms prior to hospital admission was 2 days (IQR:1-4). **Table 1** presents the characteristics of pregnant women with confirmed SARS-CoV-2 infection.

**Table 1:** Characteristics of pregnant women with confirmed SARS-CoV-2 infection

| **Characteristic** | **Total number = 185 (%)** |
| --- | --- |
| Age, years (median, IQR) | 31 (27.5-34) |
| Parity: |  |
| Nulliparous | 36 (19.5%) |
| Multiparous | 149 (80.5%) |
| Pre-existing medical disease: |  |
| Hypothyroidism | 7 (3.9%) |
| Bronchial Asthma | 4 (2.2%) |
| Chronic diabetes | 1 (0.5%) |
| Others (rheumatoid arthritis n=1, anxiety n=1, peptic ulcer=1, n=1, Crohn's disease, n=1, beta-thalassemia minor=1) | 5 (2.8%) |
| Missing data | 5 (2.8%) |
| Gestational diabetes | 19 (10.3%) |
| Preeclampsia | 4 (2.3%) |
| Gestational Age at diagnosis, weeks (median, IQR) | 29 (18-34) |
| Trimester: |  |
| 1<sup>st</sup> (less than 12 weeks) | 21 (11.4%) |
| 2<sup>nd</sup> (13-27 weeks) | 64 (34.6%) |
| 3<sup>rd</sup> (> 28 weeks) | 95 (51.3%) |
| Postpartum | 5 (2.7%) |
| Multiple pregnancies | 2 (1.1%) |
| Symptoms: | 165 |
| Fever | 105 (58%) |
| Cough | 90 (50.6%) |
| Loss of smell | 10 (5.8%) |
| Sore throat /rhinorrhoea | 42 (24.3%) |
| Malaise/Fatigue | 26 (15.1%) |
| Chest pain/shortness of breath | 22 (12.8%) |
| Vomiting/diarrhea | 11 (6.4%) |
| Duration of symptoms prior to admission, days | 2 (1-4) |
| Asymptomatic | 21 (11.3%) |

The laboratory findings of pregnant women are shown in **Table 2**. Seventeen (9.2%) of the pregnant patients had leukopenia, and 29 (15.7%) had lymphopenia. Forty-six (37%) patients had elevated C-reactive protein and 12 (11%) had elevated procalcitonin. Elevated alanine aminotransferase (ALT), aspartate aminotransferase (AST), D-dimer and lactate dehydrogenase (LD) were observed in 24.2%, 11.2, 10% and 42%, respectively of the patients in which the test was ordered. Eight (15.4%) patients had a chest x-ray diagnosis of pneumonia.

**Table 2:** Laboratory and radiological characteristics of pregnant women

| Laboratory test | Laboratory value |  |
| --- | --- | --- |
|  | N | Median (IQR)<br>N (%) |
| White blood count (WBC) $\times 10^9$ per L<br>WBC Less than 4 | 185 | 6.7 (5.6-9.2)<br>17 (9.2%) |
| Lymphocytes $\times 10^9$ /L<br>Lymphocyte less than 1<br>Lymphocyte more than 3.6 | 185 | 1.5 (1.1-2.05)<br>29 (15.7%)<br>32 (17.3%) |
| Neutrophils $\times 10^9$<br>Neutrophils less than 1.8<br>more than 7.1 | 185 | 4.8 (3.5-6.8)<br>14 (7.6%)<br>64 (34.6%) |
| Platelets $\times 10^9$<br>Platelets Less than 100 | 185 | 214 (178-252)<br>2 (1.1%) |
| C-reactive protein (CRP) (mg/L)<br>CRP> 8 | 124 | 4.5 (1.1-14.6)<br>46 (37.1%) |
| Procalcitonin (PCT) (ng/mL)<br>PCT>0.2 | 108 | 0.05 (0.05-0.08)<br>12 (11.1%) |
| Aspartate aminotransferase (AST) (U/L)<br>AST more than 40 | 125 | 21 (16-28)<br>14 (11.2%) |
| Alanine aminotransferase (ALT) (U/L)<br>ALT more than 40 | 149 | 13 (11-24)<br>36 (24.2%) |
| Lactate dehydrogenase (LDH) (IU/L)<br>LDH more>200 | 121 | 181 (154-222)<br>51 (42.1%) |
| D-Dimer (ng/mL)<br>500-1000<br>>1500 | 100 | 488.5 (333-754.5)<br>6 (6%)<br>4 (4%) |
| Albumin (g/L)<br>Albumin less than 30 | 148 | 32 (30-35)<br>31 (20.9%) |
| Chest x-ray diagnosis of pneumonia | 52 | 8 (15.4%) |

Maternal treatment and outcomes are shown in **Table 3**. Two (1.1%) patients required critical care for worsening respiratory conditions. The majority (97.3%) of the patients received Oseltamivir, and only 2 (1.1%) received Lopinavir-ritonavir. Ceftriaxone was given in 24.3% of the patients, and 96.7% received low molecular weight heparin.

**Table 3:** Maternal hospital treatment and outcomes

| **Characteristics** | **Number (%) of patients** |
| --- | --- |
| Drug therapy: |  |
| Oseltamivir | 180 (97.3%) |
| Lopinavir-ritonavir | 2 (1.1%) |
| Ceftriaxone | 45 (24.3%) |
| Azithromycin | 5 (2.7%) |
| low molecular weight heparin | 179 (96.7%) |
| Antenatal steroid for lung maturation | 38 (20.5%) |
| Intensive care unit (ICU) admission | 2 (1.1%) |
| ECMO | 0 |
| Final outcomes: |  |
| Died | 0 |
| Discharged | 183 (98.9%) |
| Still admitted | 2 (1.1%) |
| Length of hospital stay, days (median, IQR) | 15 (14-22) |

ECMO; extracorporeal membrane oxygenation

Two patients requiring Intensive care unit (ICU) were at 37 and 38 weeks’ gestation. One developed hypoxia (oxygen saturation, 88%) and shortness of breath due to worsening pneumonia necessitating intubation and emergency C-section, while the second patient had worsening respiratory condition 24 hours post-delivery **(Table S1)**. None of them required extracorporeal membrane oxygenation (ECMO), and both got discharged subsequently. At the time of follow-up, most of the patients (98.9%) were discharged, and the median duration of hospital stay was 15 days (IQR:14-22).

Pregnancy and neonatal outcomes are shown in **Table 4**. At the time of the analysis, out of the 185, 141 (76.2%) of the women had ongoing pregnancies, 40 (21.6%) gave live birth, 3 (1.6%) had a miscarriage, and 1 (0.54%) had intrauterine fetal death, which was not related to COVID-Details of the cases resulting in spontaneous miscarriages are shown in **Tables S2**. Among those who continued their pregnancy and got discharged 55 (39%) women were followed up by either telehealth or in-person at the feto-maternal clinic at median weeks post-diagnosis 3 [2-6 weeks], 25 (45.4%) patients in their 3^rd^ trimester, 17 (31%) in 2^nd^ trimester, and 13 (23.6%) in the first trimester. All these patients had a healthy pregnancy.

**Table 4:** Pregnancy and neonatal outcomes

| **Outcomes** | **Number (%) of patients** |
| --- | --- |
| Pregnancy outcomes: |  |
| Continued pregnancy | 141 (76.2%) |
| Miscarriage | 3 (1.6%) |
| Intrauterine fetal death (IUFD) | 1 (0.54%) |
| Delivered live birth (including one twin birth) | 40 (21.6 %) |
| Gestational age at delivery, weeks |  |
| Median (interquartile range) | 38 (36-39) |
| 30-32<sup>+6</sup> weeks | 2 (4.9%) |
| 33-36<sup>+6</sup> weeks | 9 (22%) |
| ≥ 37 weeks | 30 (73.1%) |
| Mode of delivery, N= 40 |  |
| C-section | 17 (42.5%) |
| Elective (repeat/maternal request) | 10 (58.8%) |
| Emergency | 7 (41.2%) |
| Maternal Hypoxia |  |
| Preeclampsia | 1 (14.3%) |
| Failed induction/ failure to progress | 1 (14.3%) |
| Fetal distress | 3 (42.8%) |
| Type of anesthesia during C-section | 2 (11.8%) |
| • General |  |
| • Spinal | 2 (11.8%) |
|  | 15 (88.2%) |
| Vaginal delivery |  |
| Instrumental delivery | 23 (57.5%) |
|  | 0 |
| Prolonged rupture of membrane (PROM>18 hours) | 3 (7.3%) |
| Birth weight (grams), median (IQR) | 3006 (2750-3215) |
| Head circumference (cm), median (IQR) | 33 (32-34.5) |
| 1 minute Apgar score, mean ±(SD) | 8 (1) |
| 5 minute Apgar score, mean ±(SD) | 9 (1) |
| Neonatal death | 0 |
| Positive for COVID-19 |  |
| None | 39 (95.1%) |
| Day 5 swab positive test | 2 (4.9%) |
| Day 14 swab positive | 1 (2.4%) |

The median gestational age at delivery was 38 (IQR: 36-39) weeks. Among those who delivered, 11(26.8%) had preterm birth. One of the preterm deliveries was iatrogenic (due to pathological cardiotocography) and the second preterm delivery due to premature rupture of membrane. Nine patients delivered between 33 and 36 weeks, either spontaneous delivery or due to other obstetric causes.

Forty-one neonates were delivered with one set of twins. Seventeen (42.5%) of neonates were born by cesarean section. Of these, one of the neonates was delivered by emergency cesarean due to maternal hypoxia as a result of worsening pneumonia, and the other 16 were due to either fetal distress or obstetrical indication. Median birth weight was 3006 grams (IQR:2750-3215), and only 2 (4.9%) neonates required intubation and surfactant due to hyaline membrane disease of prematurity. Two (4.9%) of the neonates tested positive for SARS-CoV-2 infection, and there was no neonatal death.

Clinical characteristics of SARS-CoV-2 positive neonates are shown in **Table S3**. The first positive neonate was born at 31 weeks by vaginal delivery; the mother presented with preterm premature rupture of membrane and developed intrapartum fever (38.2^0^). The obstetrical cause of fever was ruled out, and the COVID-19 was suspected that was later confirmed on RT-PCR. The second positive neonate was born at 39 weeks’ gestation by spontaneous vaginal delivery, similar to other positive newborns; the mother also had premature rupture of membrane. One newborn was asymptomatic apart from being born prematurely at 31 weeks. The second newborn developed tachypnoea and increase work of breathing on day 4 of age and required humidified high flow nasal cannula for 5 days. The neonate’s clinical condition improved, and a chest x-ray showed complete resolution of the bilateral streaky lung infiltration.

## Discussion

This is a descriptive study on the maternal as well as neonatal clinical features and outcome of pregnancies complicated with SARS-CoV-2 infection. Thus far, the majority of the preliminary research articles reported with a small sample size [12, 13, 15, 18]. This study is among the first to report COVID-positive maternal and neonatal clinical characteristics and outcomes with a comparatively large sample size.

In this study, 185 pregnant women were identified with confirmed SARS-CoV-2 infection with. Around 88% were symptomatic, with the most common symptom being fever in 58% of patients followed by cough in 50.6% of patients. These findings are in accordance with a number of preliminary studies [14, 15, 18, 29]. A prospective cohort study using the UK Obstetric Surveillance System (UKOSS) found fever and cough as common symptoms in pregnant women having COVID-19 [29]. A systematic review revealed that the most dominant initial symptoms in pregnant women with COVID-19 were fever and cough [18]. Similarly, a retrospective study found that common symptoms in pregnant women in the wake of SARS-CoV-2 infection are cough and fever, and less common symptoms include shortness of breath, diarrhea, and myalgia [13]. On the contrary, a study found the majority of women being asymptomatic and afebrile at presentation [17]. However, the majority of the studies have supported the evidence of fever and cough as the most common symptoms [14, 15, 30 – 35]

Data from the preliminary studies found that majority of pregnant women with COVID-19 were in the late second or third trimester [29]. In our study, the median gestational age on diagnosis was 29 weeks, and half of the women were in their 3^rd^ trimester. Likewise, a prospective cohort study using UKOSS found that most women were hospitalized in the 3^rd^ trimester [8, 29].

Moreover, in our study, 10% of the patients had gestational diabetes. Evidence of an association between gestational diabetes and pregnant women infected with SARS-CoV-2 is well documented [29].

With reference to the laboratory findings, in our study, 15.7% of the pregnant patients had lymphopenia, and elevated ALT and AST were observed in 24.2% and 11.2% of the patients, respectively, and 10% had elevated D-dimer. Several studies have reported lymphopenia in both the general population affected by COVID-19 [36] and also in pregnant women affected by COVID-19 [17, 37]. Similarly, a retrospective study found the likelihood of lymphopenia as well as increased concentrations of ALT or AST as one of the clinical manifestations [13]. Pregnancy is widely accepted as a hypercoagulable state [38], and monitoring of D-dimer is also necessary. A case series study has reported elevated D-dimer in pregnant women with COVID-19 and its association with increased mortality rate [39].

Preliminary studies on pregnancies complicated with SARS-CoV-2 infection suggested a low rate of admission to the ICU, maternal, and neonatal death [40]. Moreover, the rate of severe pneumonia in pregnant women was also not greater than the general population [41]. In our study, 15.4% had a radiological diagnosis of pneumonia, and only 2 patients required critical care for worsening respiratory conditions, which are in accordance with the preliminary records. A case series published by the clinicians in New York reported a similarly low rate of around 5% of the pregnant patients with SARS-CoV-2 infection requiring critical care [14]. Likewise, a prospective cohort study using UKOSS reported that one woman per 2400 giving birth required critical care, and the maternal mortality rate was roughly about one in 18000 women undergoing delivery [29].

The rates of ICU admission and mortality among pregnant women admitted to hospital with COVID-19 are proportional to the rates among the general population [29, 41]. In our study, we found favorable outcomes among pregnant women with SARS-CoV-2 infection. Most of the patients had mild symptoms, with only a few ICU admissions and zero mortality. The clinical features of pregnant women with SARS-CoV-2 infection were similar to those of the general population having SARS-CoV-2 infection, as reported in previous studies [42].

In our study, 26.8% of neonates were born preterm, and 42.5% of neonates were delivered by cesarean section. These findings are supported by similar findings in the past, in the data from the UKOSS study, where 59% of the neonates had cesarean births, and 27% had preterm births [29]. Another study reported that the majority of pregnant women had a cesarean section planned delivery to prevent neonatal transmission of the virus [34]. Similarly, a systematic review finds out that the frequent mode of delivery in cases reported was cesarean section [18].

Newborns born to SARS-CoV-2 infected mothers are most likely to be asymptomatic [12]. Similarly, in our study, 73.1% of the neonates, born full-term, were asymptomatic. The findings of our study propose that neonatal outcome is reassuring, keeping with other studies [29, 43-46]. However, in our study, two neonates tested positive for COVID-19 on day 5 of nasopharyngeal swab testing. According to Shah’s classification, the diagnosis of congenital or intrapartum infection could not be confirmed [47]. Some studies from the past reported inconsistency with no evidence of vertical transmission [13, 34]. Nevertheless, vertically acquired infection is still reasonable and cannot be ruled out as supported by preliminary studies and emerging evidence [8]. Further investigations around the vertical transmission of SARS-CoV-2 infections is needed.

In our study, the mothers of two COVID-19-positive neonates presented with preterm premature rupture of membrane (PROM). PROM is known to be linked to neonatal sepsis, and duration of PROM and viral infection in neonate, especially with herpes simplex and HIV, is well documented, however, it is difficult to make a definitive conclusion here [48, 49]

A major strength of this study is its large sample size, whereas preliminary studies [12, 15, 18] were comprised of small sample size and could not draw a firm conclusion. The findings of the study may aid clinicians and patients in the effective management of the pregnancies complicated with SARS-CoV-2 infection. However, there are also some inherent limitations to this study due to its retrospective study design.

In conclusion, the clinical features of pregnant women with SARS-CoV-2 infection were similar to those of the general population infected with COVID-19. Favorable outcomes among pregnant women with SARS-CoV-2 infection suggest that pregnant women are not at higher risk to the COVID 19 than the general population, nor are they susceptible to severe pneumonia. Moreover, the neonatal outcome is also reassuring.

## Data Availability

Available

## Supporting information

Appendix 1. Management of Patients with COVID-19 (PDF)

### Supplementary file

**Table S1.** Clinical features of pregnant patients with COVID-19 in ICU

|  | **Patient 1** | **Patient 2** |
| --- | --- | --- |
| Age | 33 | 39 |
| Parity | 0 | 1 |
| Comorbidities | GDM on insulin | Healthy |
| Presenting symptoms | Fever for 2 days | Fever and cough for 4 days |
| Gestational age at diagnosis, weeks | 38+2 | 37 |
| Mode of delivery | Emergency C/S (failure to progress) | Emergency c/s due to maternal hypoxemia |
| Time of ICU admission | 24 hours after delivery | Immediately post-delivery |
| Cause of ICU admission | Respiratory distress | Hypoxemia |
| ARDS-Berlin score | Moderate | Moderate |
| qSOFA | 2 | 3 |
| Maximum respiratory support | Intubated | Intubated |
| ECMO | 0 | 0 |
| Laboratory findings (on ICU admission) |  |  |
| White blood cell x 10<sup>9</sup> | 4.8 | 18.4 |
| Lymphocyte x 10<sup>9</sup> | 0.7 | 1.1 |
| Neutrophil x10<sup>9</sup> | 3.6 | 16.4 |
| Hemoglobin (g/dl) | 123 | 115 |
| Platelets x10<sup>9</sup> | 118 | 393 |
| C-reactive protein (mg/L) | 200 | 19.6 |
| D-Dimer (ng/mL) | 774 | 906 |
| Alanine transaminase (U/L) | 14 | 11 |
| Aspartate transaminase (U/L) | 36 | 31 |
| Lactate dehydrogenase (IU/L) | 552 | 356 |
| Duration of intubation, days | 3 | 6 |
| Duration of ICU stay, days | 5 | 12 |
| Duration of hospitalization, days | 15 | 20 |
| ARDS: Acute respiratory distress syndrome, qSOFA: quick Sequential Failure Assessment, ECMO: extracorporeal membrane oxygenation, GDM: Gestational Diabetes Mellitus<br>ARDS-Berlin score: Acute respiratory distress syndrome (ARDS) is classified into three stages i.e., mild, moderate, and severe, according to oxygenation severity at the onset. |  |  |

**Table S2.** Clinical features of pregnant COVID-19 patients who had a miscarriage

|  | **Patient 1** | **Patient 2** | **Patient 3** |
| --- | --- | --- | --- |
| Maternal age, years | 37 | 29 | 31 |
| Parity | 1 | 1 | 2 |
| Past Obstetrical history | 0 | 0 | 0 |
| Pre-existing comorbidities | 0 | 0 | 0 |
| Gestational Diabetes | 0 | 0 | 0 |
| PIH | 0 | 0 | 0 |
| Outcome | Miscarriage | Miscarriage | Miscarriage |
| Gestational age at diagnosis, weeks | 14 | 13 | 14 |
| Symptoms at diagnosis | Sore throat<br>Malaise/fatigue | Runny nose | Fever<br>Cough |
| Duration of symptoms prior to diagnosis | 5 days | 1 day | 2 days |
| Total WBC x 10<sup>9</sup> | 2.3 | 6.5 | 4.5 |
| Neutrophil x 10<sup>9</sup> | 0.67 | 4.1 | 3.4 |
| Lymphocytes x 10<sup>9</sup> | 1.5 | 1.4 | 0.8 |
| Hemoglobin (g/L) | 82 | 120 | 133 |
| Platelets x 10<sup>9</sup> | 67 | 212 | 287 |
| LDH (IU/L) | Not available | 112 | 181 |
| CRP (mg/dl) | 48 | 22 | 75 |
| INR, ratio | 1.2 | 1.27 | 1.25 |
| PT, seconds | 17.8 | 17.6 | 17 |
| D-Dimer (ng/ml) | Not available | 452 | 532 |
| ALT (U/L) | 14 | 22 | 11 |
| ALP (U/L) | 68 | 39 | 39 |
| Albumin (g/L) | 39 | 39 | 35 |
| PIH: Pregnancy-induced hypertension, Total WBC: Total white blood cells, LDH: Lactate Dehydrogenase, CRP: C-Reactive Protein, INR: International Normalized Ratio, PT: Prothrombin Time, ALP: Alkaline Phosphatase |  |  |  |

**Table S3.**
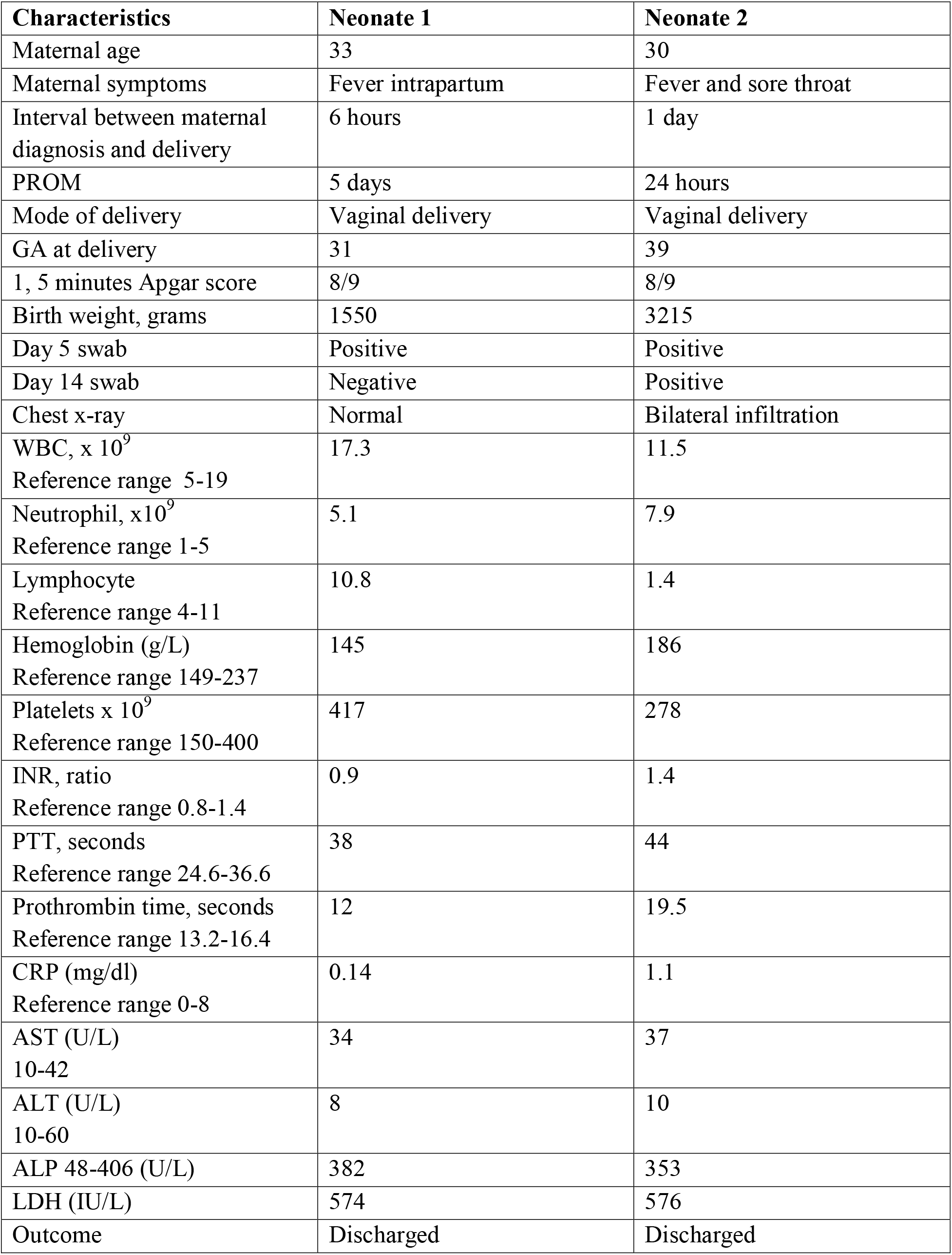
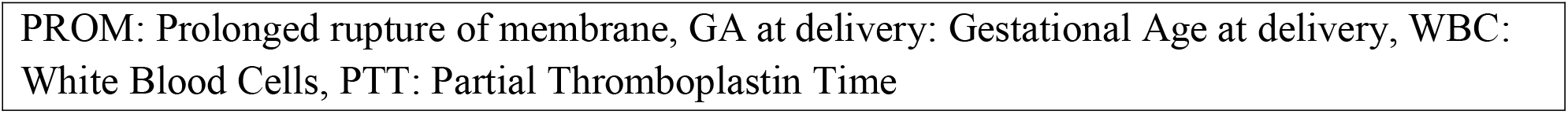
Clinical features of COVID-19 positive neonate

| **Characteristics** | **Neonate 1** | **Neonate 2** |
| --- | --- | --- |
| Maternal age | 33 | 30 |
| Maternal symptoms | Fever intrapartum | Fever and sore throat |
| Interval between maternal diagnosis and delivery | 6 hours | 1 day |
| PROM | 5 days | 24 hours |
| Mode of delivery | Vaginal delivery | Vaginal delivery |
| GA at delivery | 31 | 39 |
| 1, 5 minutes Apgar score | 8/9 | 8/9 |
| Birth weight, grams | 1550 | 3215 |
| Day 5 swab | Positive | Positive |
| Day 14 swab | Negative | Positive |
| Chest x-ray | Normal | Bilateral infiltration |
| WBC, $\times 10^9$<br>Reference range 5-19 | 17.3 | 11.5 |
| Neutrophil, $\times 10^9$<br>Reference range 1-5 | 5.1 | 7.9 |
| Lymphocyte<br>Reference range 4-11 | 10.8 | 1.4 |
| Hemoglobin (g/L)<br>Reference range 149-237 | 145 | 186 |
| Platelets $\times 10^9$<br>Reference range 150-400 | 417 | 278 |
| INR, ratio<br>Reference range 0.8-1.4 | 0.9 | 1.4 |
| PTT, seconds<br>Reference range 24.6-36.6 | 38 | 44 |
| Prothrombin time, seconds<br>Reference range 13.2-16.4 | 12 | 19.5 |
| CRP (mg/dl)<br>Reference range 0-8 | 0.14 | 1.1 |
| AST (U/L)<br>10-42 | 34 | 37 |
| ALT (U/L)<br>10-60 | 8 | 10 |
| ALP 48-406 (U/L) | 382 | 353 |
| LDH (IU/L) | 574 | 576 |
| Outcome | Discharged | Discharged |

PROM: Prolonged rupture of membrane, GA at delivery: Gestational Age at delivery, WBC: White Blood Cells, PTT: Partial Thromboplastin Time

## Notes

### Author Declarations

The study was approved by the Ethics Committee of the Ministry of Health of Kuwait (2020/1420)

